# Multimodal magnetic resonance imaging characterizes clinical outcome in chronic traumatic brain injury

**DOI:** 10.1101/2024.08.10.24305340

**Authors:** Mélanie Pélégrini-Issac, Adrienne Hezghia, Elsa Caron, Sébastien Delphine, Valentine Battisti, Didier Cassereau, Clara Debarle, Muriel Lefort, Blandine Lesimple, Grégory Torkomian, Vincent Degos, Rémy Bernard, Damien Galanaud, Pascale Pradat-Diehl, Vincent Perlbarg, Eléonore Bayen, Louis Puybasset

## Abstract

Moderate to severe traumatic brain injury (TBI) should be considered as a chronic health condition. The corpus callosum is the brain region that suffers most from diffuse axonal injury, leading to long-term functional deficits. Few studies have considered the relationships between inter- and intrahemispheric functional connectivity and structural damages to the corpus callosum in chronic TBI patients. We examined how callosal functional connectivity and white matter alterations relate to clinical outcome using multimodal magnetic resonance imaging (MRI): structural MRI estimates callosal volume, diffusion-weighted MRI enables white matter integrity quantification, resting-state functional MRI assesses neural dysfunction. Seventy-four patients underwent a multimodal MRI session on average 5 years after a moderate-to-severe TBI. Multiple factorial analysis analyzed the relationships between clinical outcome (from severe disability to good recovery, assessed by the Glasgow Outcome Scale extended GOSE), callosal volume, diffusion metrics (fractional anisotropy and mean, axial, and radial diffusivity), and inter- and intrahemispheric functional connectivity. Multiple factorial analysis confirmed that patients with severe disability (GOSE 3-4) had more structural alterations in the corpus callosum than patients with a good recovery (GOSE 7-8). Most importantly, patients able to live independently but unable to work/study in a standard environment (GOSE 5-6) could not be described solely by structural features. They exhibited a lower interhemispheric connectivity between cortical regions mediated by the corpus callosum than patients with a good recovery, and a tendency towards a decrease in intrahemispheric connectivity compared with severely disabled patients. These findings suggest a complex long-term functional impact of moderate-to-severe TBI.

## 1. Introduction

Traumatic brain injury (TBI) is a significant global public health concern referred to as a “silent epidemic” expected to be one of the top three causes of disability worldwide [1]. Due to advancements in health care and biotechnology, more patients are surviving a moderate to severe TBI (defined by a loss of consciousness of 30 minutes or more, and a post-traumatic anterograde amnesia of 24 hours or more [2]) that would have previously resulted in death. Nevertheless, many of them experience long-term disability, characterized by a variety of physical, cognitive, emotional, and behavioral difficulties.

Although focal lesions such as contusions and intracranial hemorrhage may occur in severe TBI, widespread microscopic lesions to white matter tracts, known as diffuse axonal injury, are most common: Axons experience strong shearing forces that cause them to tear from the cell body, resulting in neuronal death [3]. This can disrupt neural communication in cortical and subcortical networks, leading to a broad range of functional deficits that may last for years beyond the initial injury. Increasing evidence suggests that TBI is a progressive disorder that can lead to greater neurodegeneration over time and should be conceptualized as a chronic health condition [4,5]. Long-term disability is usually assessed by the eight-point extended Glasgow Outcome Scale (GOSE), which reflects a patient’s overall social outcome and his/her capacity in domains such as dependence on others and social reintegration [6]. GOSE scores have been found to remain fairly constant in moderate-to-severe TBI patients over the first five years after injury [7].

Detecting white matter disruptions in the living brain is essential to understand long-term functional impairments of TBI patients. However, conventional diagnostic neuroimaging including computed tomography and structural magnetic resonance imaging (MRI) fails to reliably detect diffuse axonal injury in vivo. Combined with the tensor model (the so-called diffusion tensor imaging [DTI]), diffusion-weighted imaging has come to represent the modality of reference to quantify white matter integrity, giving access to local diffusion metrics including fractional anisotropy (FA) and mean diffusivity (MD) [8]. Resting-state functional MRI (rs-fMRI) can further assess patterns of network dysfunction induced by TBI by measuring spontaneous fluctuations in low-frequency (0.005 – 0.08 Hz) blood-oxygen-level dependent (BOLD) signal in the absence of an external stimulus [9]. Significant temporal correlations between BOLD signals from spatially distinct brain regions reflect functional connectivity [10]. Several studies have shown functional connectivity changes after brain injury [11,12].

As the largest white matter bundle in the brain and a stable structure attached to the large and comparatively more moveable cerebral hemispheres, the corpus callosum is a critical region that is one of the most susceptible to diffuse axonal injury [13]. The corpus callosum plays a major functional role in both excitatory and inhibitory interhemispheric communication between homologous regions by exchanging sensory, cognitive, and motor information [8]. Task-based functional MRI studies have related callosal white matter alterations following TBI to functional and cognitive deficits such as impaired bimanual coordination and verbal and visuospatial working memory [14,15]. Besides, functional connectivity rs-fMRI studies have led to controversial results regarding an increase or a decrease in connectivity following TBI and only few studies have considered the possible relationship between hemispheric functional connectivity and structural damages to the corpus callosum and its subregions (genu/anterior, body/central and splenium/posterior) as measured by DTI metrics [16,17]. Such diversity may be attributed to the fact that sample sizes have remained rather small (typically less than 20-30 patients) and heterogeneous without any stratification in terms of patient severity (some studies mixing mild, moderate, and severe TBI patients [18]) or long-term disability (based on the GOSE score), and with variable follow-up durations (from a few months to several years after the injury [19,20]).

To better understand which brain imaging features may help distinguish chronic moderate-to-severe TBI patients in terms of recovery (from severe disability to good recovery), the objective of the present retrospective study is to provide a statistical description, using multivariate factor analysis, of how functional connectivity patterns and white matter integrity alterations in the corpus callosum relate to clinical outcome as assesseed by the GOSE.

## 2. Methods

### 2.1. Participants

This descriptive study is part of a large data collection project on the long-term follow-up of patients with severe TBI. Patient management was not modified. In accordance to French law, this retrospective and monocentric study was approved by the ethics committee “Comité éthique pour la recherche en anesthésie-réanimation”, Paris, France (IRB 00010254-2024-007). All patients expressed non opposition to participating in this study after receiving written information. Adult patients were recruited at least two years after the injury between November 2012 and December 2016. During clinical evaluation, EB, EC, CD and BL had access to information that could identify the participants. All data were then pseudonymized prior to analysis.

Inclusion criteria were (1) at least 18 years old at the time of GOSE assessment, (2) worst Glasgow Coma Scale (GCS) score of 3-15 at the time of injury, (3) injury requiring hospitalization in a neurosurgical intensive care unit, (4) follow-up at least 2 years post injury, and (5) fluency in French. Exclusion criteria included (1) trauma caused by a ballistic brain injury, (2) pre-existing neurological disorders, (3) history of psychiatric disorders, (4) pre-existing addiction to cannabis, heroin, or cocaine, (5) addiction to alcohol combined with either another drug, or with major depressive disorder or epilepsy, (6) drug replacement therapy, (7) sensory impairment preventing neuropsychological assessment (e.g. blindness, deafness), (8) contraindication to MRI, (9) physical inability to come to the hospital for a one-day exam, and (10) absence of complete refund of medical exams by the French National Health Insurance.

### 2.2. Clinical evaluation

Clinical data included patient characteristics, initial clinical status including the worst GCS score measured at the time of injury, and GOSE score at the time of participation. GOSE classifies clinical outcome into eight categories including: dead (GOSE = 1), vegetative state (GOSE = 2), lower severe disability (GOSE = 3), upper severe disability (GOSE = 4), lower moderate disability (GOSE = 5), upper moderate disability (GOSE = 6), lower good recovery (GOSE = 7), and upper good recovery (GOSE = 8) [6]. Patients were divided into three groups based on their GOSE score: Patients with a GOSE score of 3-4 were categorized as “disabled” (able to follow commands but need someone else’s help for daily living activities), patients with a score of 5-6 were categorized as “intermediate” (able to live independently but unable to work/study in a standard environment), and patients with a score of 7-8 were categorized as “good recovery” (able to return to work/school but presenting with cognitive complaints or minor behavioral deficits that impair the quality of their everyday life with family and friends, and interfere with their participation in social and leisure activities).

### 2.3. MRI metrics

MRI reporting guidelines from the Committee on Best Practices in Data Analysis and Sharing (COBIDAS) were followed [21]. Data acquisition protocol and preprocessing methods are provided in supplemental material.

#### 2.3.1. Structural data

After preprocessing the three-dimensional structural MRI data, the corpus callosum was segmented into three regions of interest (the genu, the body and the splenium). All volume values were expressed as a proportion of the estimated total intracranial volume.

#### 2.3.2. DTI data

After preprocessing the diffusion-weighted MRI data, the values of the four diffusion metric maps (fractional anisotropy [FA], mean diffusivity [MD], diffusion along the main axis [axial diffusivity AD], and diffusion along the transverse plane [radial diffusivity RD]) were averaged across voxels of the genu, the body and the splenium of the corpus callosum defined from the ICBM-DTI-81white matter atlas [22].

#### 2.3.3. Resting-state functional MRI data

After preprocessing the rs-fMRI data, BOLD signal time courses were averaged within each of the 82 cortical parcels of the Automated Anatomical Labeling (AAL) atlas [23]. Functional connectivity was assessed by calculating Pearson’s correlation coefficient *ρ*(*R*_*i*_, *R*_*j*_) between the average time courses of all possible pairs of regions (*R*_*i*_ and *R*_*j*_, where *i* ≠ *j*).

AAL labels were divided into three subgroups in relation to their interhemispheric white matter connections through the corpus callosum, in line with Hofer and Frahm’s scheme (Table 1) [24]. Interhemispheric and intrahemispheric functional connectivity measures were computed separately for each subgroup of regions connected by a given subdivision of the corpus callosum, as described in Appendix 1.

**Table 1.** AAL atlas regions associated with subdivisions of the corpus callosum.

| **Genu** | **Body** | **Splenium** |
| --- | --- | --- |
| 1 Superior frontal gyrus, dorsolateral | 1 Precentral gyrus | 1 Posterior cingulated gyrus |
| 2 Superior frontal gyrus, orbital part | 2 Rolandic operculum | 2 Hippocampus |
| 3 Middle frontal gyrus | 3 Supplementary motor area | 3 Parahippocampal gyrus |
| 4 Middle frontal gyrus, orbital part | 4 Olfactory cortex | 4 Amygdala |
| 5 Inferior frontal gyrus, opercular part | 5 Median cingulate and paracingulate gyri | 5 Calcarine fissure and surrounding cortex |
| 6 Inferior frontal gyrus, triangular part | 6 Paracentral lobule | 6 Cuneus cortex |
| 7 Inferior frontal gyrus, orbital part |  | 7 Lingual gyrus |
| 8 Superior frontal gyrus, medial |  | 8 Superior occipital gyrus |
| 9 Medial orbitofrontal cortex |  | 9 Middle occipital gyrus |
| 10 Gyrus rectus |  | 10 Inferior occipital gyrus |
| 11 Insula |  | 11 Fusiform gyrus |
| 12 Anterior cingulate gyrus |  | 12 Postcentral gyrus |
|  |  | 13 Superior parietal gyrus |
|  |  | 14 Inferior parietal, excluding supramarginal and angular gyri |
|  |  | 15 Supramarginal gyrus |
|  |  | 16 Angular gyrus |
|  |  | 17 Precuneus |
|  |  | 18 Heschl gyrus |
|  |  | 19 Superior temporal gyrus |
|  |  | 20 Temporal pole, superior temporal gyrus |
|  |  | 21 Middle temporal gyrus |
|  |  | 22 Temporal pole, middle temporal gyrus |
|  |  | 23 Inferior temporal gyrus |

### 2.4. Statistical Analyses

Between-group differences in gender were assessed with Fisher’s exact test. Differences in age at the time of GOSE assessment and delay between injury and MRI session were examined with the Kruskal-Wallis test.

Spearman’s rank-order correlations between imaging metrics were determined. Permutational multivariate ANOVA (MANOVA, 10,000 permutations) was used to assess significant effects between groups of patients (*P*-values adjusted by controlling the false discovery rate [25]). If a significant group effect was found, a Kruskal-Wallis test was conducted on each metric (*P*-value Bonferroni-corrected by the number of tests conducted) and followed by pairwise Dunn’s tests (corrected for multiple testing using Benjamini-Hochberg’s false discovery rate procedure [26]) to assess which metric differed between which groups.

Finally, multiple factorial analysis (MFA, Appendix 2) was used to summarize and cluster information contained in the variables of interest [27]. In this study, variables were grouped as follows: clinical information (GOSE category), anatomy (callosal subregion volume), diffusion (FA, MD, AD, RD metrics), and connectivity (inter- and intrahemispheric functional connectivity measures). Each subdivision in the corpus callosum was analyzed separately.

Statistical analyses were performed using R (4.3.3) and MFA was conducted using *FactoMineR* and *factoextra* packages [28–30].

## 3. Results

As results were similar for the three subdivisions of the corpus callosum, only results obtained for regions connected by the splenium are reported. Results obtained for the body and the genu are available in supplemental material.

### 3.1. Demographics

A total of 74 patients met inclusion criteria (Table 2 and supplemental material, Table 1). Ten disabled patients (“Dis”), 42 intermediate patients (“Int”), and 22 patients with good recovery (“Rec”) participated in the study. Mechanism of injury was road traffic accident in 46 (62.2%) patients, fall in 12 (16.2%), assault in 10 (13.5%), and other causes in 6 (8.1%). All patients were in the chronic phase of TBI, with a mean time between injury and GOSE assessment and a mean delay between injury and MRI assessment of 5.2 years. There was no significant difference among patient groups in terms of age (Kruskal-Wallis *χ*^2^=0.10, *P*=0.95), initial GCS (Kruskal-Wallis *χ*^2^=5.50, *P*=0.06), gender (Fisher’s exact test *P*=0.38), and delay between injury and MRI session (Kruskal-Wallis *χ*^2^=0.10, *P*=0.95).

**Table 2.** Patients characteristics.

| **Patient group** | ***N*** | **Gender (F/M)** | **Mean age (SD)<sup>a</sup>** | **Median initial GCS (IQR)<sup>b</sup>** | **Median GOSE (IQR)** | **Mean delay to MRI (SD)<sup>c</sup>** |
| --- | --- | --- | --- | --- | --- | --- |
| **All** | **74** | **8/66** | **38.2 (15.0)** | **7 (3.5)** | **6 (2)** | **62.7 (21.7)** |
| Dis | 10 | 1/9 | 38.4 (14.6) | 6 (1) | 4 (1) | 64.3 (22.5) |
| Int | 42 | 3/39 | 38.2 (14.7) | 7 (4) | 5 (1) | 61.9 (21.6) |
| Rec | 22 | 4/18 | 38.0 (16.3) | 8 (4) | 7 (1) | 63.3 (22.5) |

<sup>a</sup>Age is at the time of GOSE assessment and is expressed in years.
<sup>b</sup>GCS was unavailable for 3 patients.
<sup>c</sup>Delay is expressed in months.
*N*: number of patients; M, male; F, female; SD: standard deviation; IQR: interquartile range; GCS, Glasgow Coma Scale score; GOSE, Glasgow Outcome Scale extended score; Dis, disabled patients; Int, intermediate patients; Rec, patients with good recovery.

### 3.2. Imaging metrics

Distributions of volume values, diffusion metrics and functional connectivity measures for the splenium of the corpus callosum are shown in Fig. 1 and Table 3. Spearman’s rank correlations between variables are shown in Table 4. The volume of the splenium was significantly correlated with all diffusion metrics, but not with functional connectivity measures. Significant correlations were found amongst diffusion metrics. In particular, RD and MD in the splenium were significantly correlated with all other metrics and were therefore dropped from further ANOVA analyses as they did not convey any additional information compared with FA and AD. No significant correlation was found between diffusion and functional connectivity metrics. Left and right intrahemispheric connectivity measures were highly significantly correlated with one another but interhemispheric connectivity only significantly correlated with the left intrahemispheric connectivity measure.

**Figure 1.**
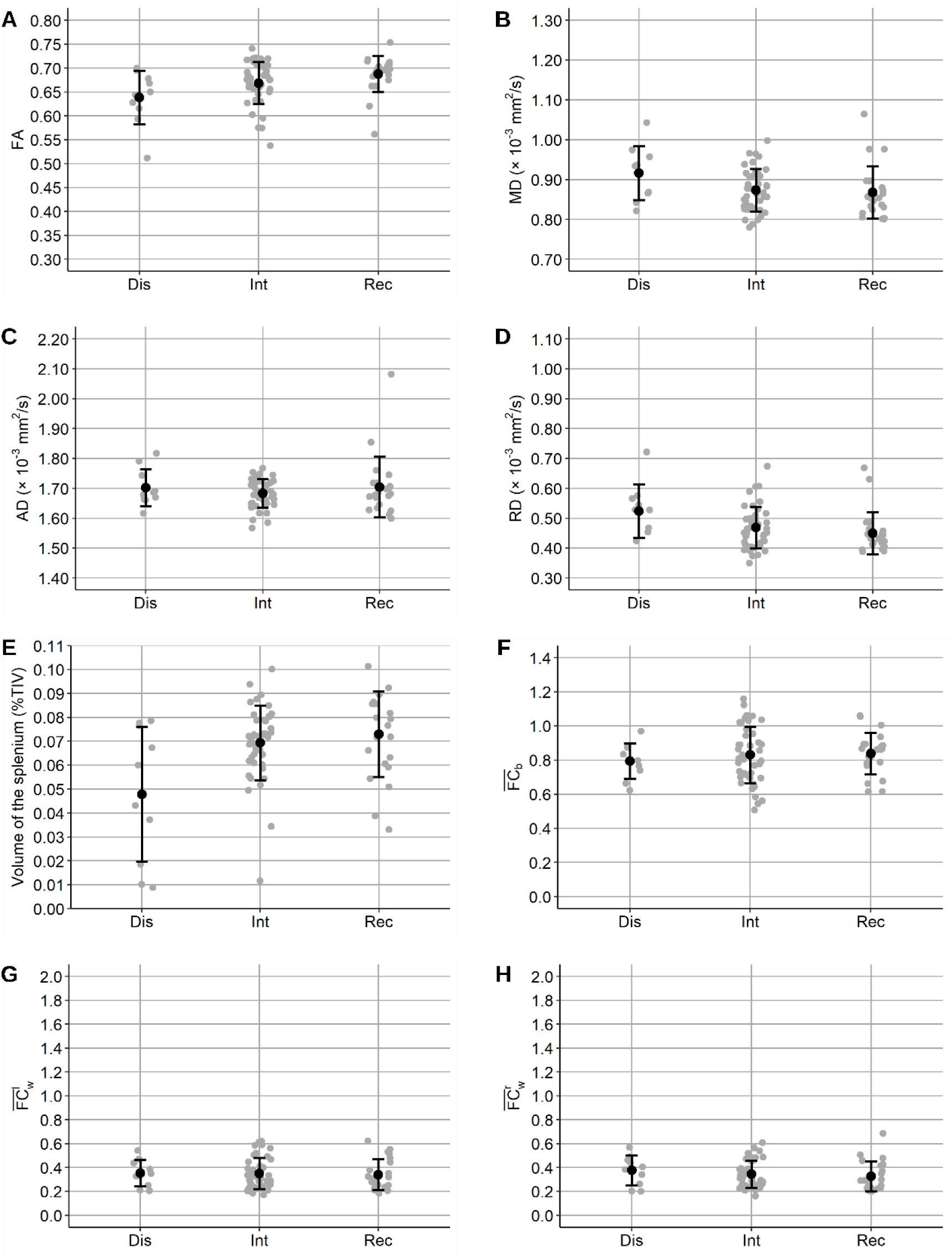
Distribution of imaging metrics for the splenium of the corpus callosum. Dis, disabled patients; Int, intermediate patients; Rec, patients with good recovery. Black dots: Smean values, vertical black lines: standard deviation, grey dots: individual values. Panels **A**: fractional anisotropy (FA), **B**: mean diffusivity (MD), **C**: axial diffusivity (AD), **D**: radial diffusivity (RD), **E**: volume, **F**: interhemispheric functional connectivity 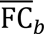, **G-H**: left 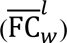 and right 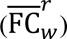 intrahemispheric functional connectivity.

**Table 3.** Splenium of the corpus callosum: Mean [standard deviation] of the imaging metrics.

| Patients | Volume<br>(%TIV) | FA | MD<br>($\times 10^{-3}$ mm<sup>2</sup>/s) | AD<br>($\times 10^{-3}$ mm<sup>2</sup>/s) | RD<br>($\times 10^{-3}$ mm<sup>2</sup>/s) | $\overline{FC}_b$ | $\overline{FC}_w^l$ | $\overline{FC}_w^r$ |
| --- | --- | --- | --- | --- | --- | --- | --- | --- |
| Dis | 0.048<br>[0.028] | 0.639<br>[0.056] | 0.916<br>[0.068] | 1.70<br>[0.063] | 0.524<br>[0.089] | 0.795<br>[0.103] | 0.352<br>[0.111] | 0.377<br>[0.124] |
| Int | 0.069<br>[0.016] | 0.669<br>[0.044] | 0.874<br>[0.053] | 1.68<br>[0.048] | 0.469<br>[0.070] | 0.830<br>[0.166] | 0.350<br>[0.130] | 0.344<br>[0.116] |
| Rec | 0.073<br>[0.018] | 0.687<br>[0.038] | 0.868<br>[0.066] | 1.70<br>[0.101] | 0.450<br>[0.071] | 0.839<br>[0.122] | 0.340<br>[0.128] | 0.327<br>[0.124] |

Dis, disabled patients; Int, intermediate patients; Rec, patients with good recovery. TIV: total intracranial volume, FA: fractional anisotropy, MD: mean diffusivity, AD: axial diffusivity, RD: radial diffusivity, $\overline{FC}_b$ : interhemispheric functional connectivity, $\overline{FC}_w^l$ : left and $\overline{FC}_w^r$ : right intrahemispheric functional connectivity.

**Table 4.** Splenium of the corpus callosum: Spearman’s rank correlation matrix between MRI measures.

| | Volume | FA | MD | AD | RD | $\overline{FC}_b$ | $\overline{FC}_w^l$ |
| --- | --- | --- | --- | --- | --- | --- | --- |
| FA | **0.41** | – | – | – | – | – | – |
| MD | **-0.46** | **-0.83** | – | – | – | – | – |
| AD | **-0.39** | -0.25 | **0.65** | – | – | – | – |
| RD | **-0.43** | **-0.97** | **0.93** | **0.43** | – | – | – |
| $\overline{FC}_b$ | 0.01 | 0.19 | -0.20 | 0.04 | -0.20 | – | – |
| $\overline{FC}_w^l$ | -0.01 | -0.01 | -0.03 | -0.06 | -0.01 | **0.42** | – |
| $\overline{FC}_w^r$ | 0.02 | -0.06 | 0.10 | 0.03 | 0.07 | 0.34 | **0.89** |

Correlations indicated in bold are significantly different from 0 ( $P < 0.05$ Bonferroni-corrected for 28 different pairwise tests, i.e. $P < 0.0018$ ).
$\overline{FC}_b$ , interhemispheric functional connectivity; $\overline{FC}_w^l$ , left intrahemispheric functional connectivity; $\overline{FC}_w^r$ , right intrahemispheric functional connectivity.

Permutational MANOVA was conducted with splenium volume, FA and AD as independent variables, showing a significant group effect (*F*=4.16, *P*=0.02). FA only was found to differ significantly (Kruskal-Wallis *χ*^2^=9.04, *P*=0.01) between “Dis” and “Rec” patients (Dunn’s test *Z*=2.91, *P*=0.01).

A separate permutational MANOVA was conducted with connectivity measures as independent variables, showing no significant difference between patient groups (*F*=0.21, *P*=0.90).

A multiple factorial analysis was conducted with variables structured into the following groups: clinical information (GOSE), anatomy (splenium volume), diffusion (FA, MD, AD and RD metrics), and connectivity (inter- and left/right intrahemispheric functional connectivity measures). The cumulative percentage of inertia explained by the first three components was 65.8%. A bar plot of group contribution to the first two dimensions is shown in Fig. 2A and Fig. 2B, showing a high contribution of anatomy and diffusion to the first dimension, and a high contribution of functional connectivity measures to the second dimension. GOSE contributed highly to both dimensions.

**Figure 2.**
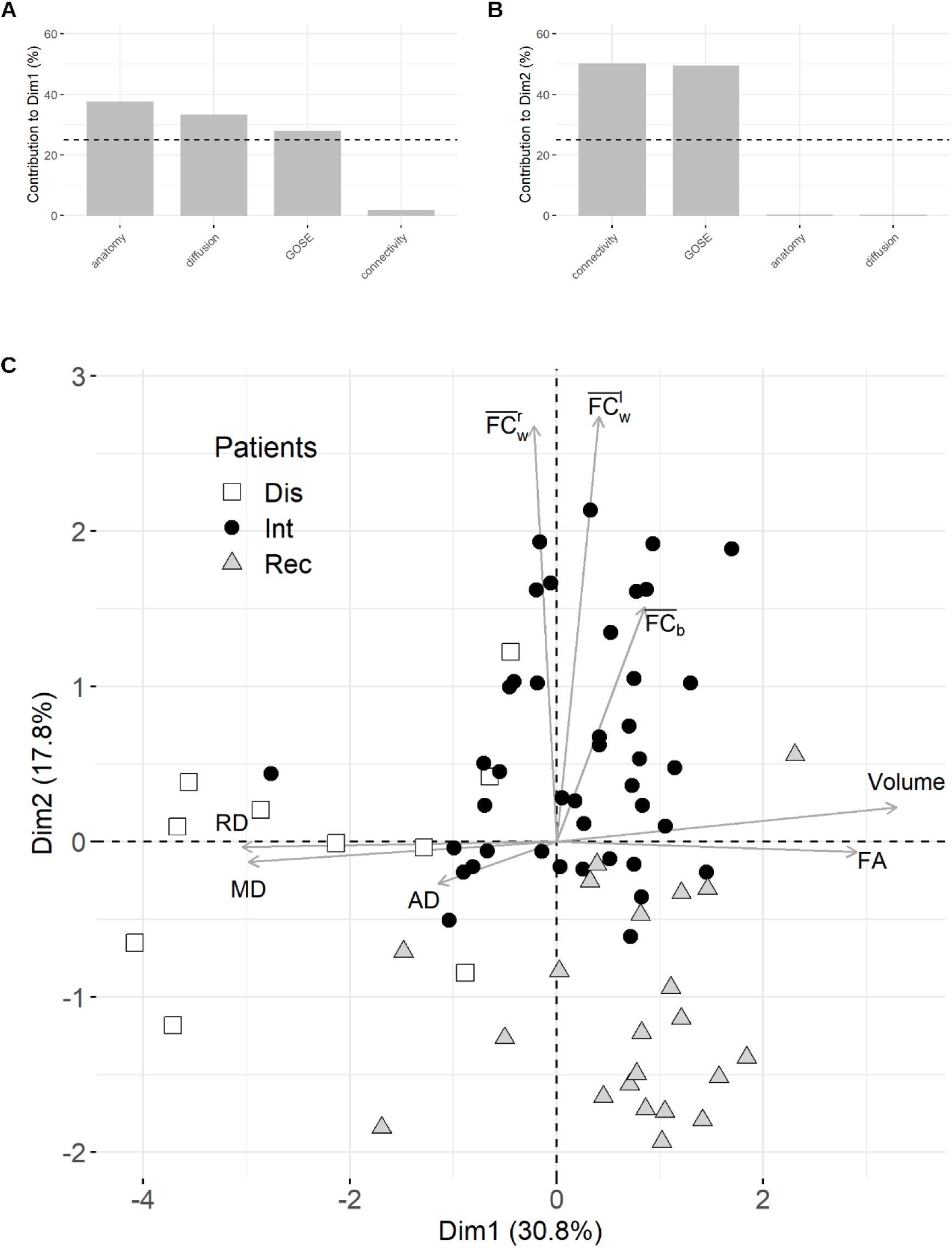
Results of the multivariate factorial analysis for the splenium of the corpus callosum. **(A-B)** contribution (in %) of each group of variables to the first two dimensions. The reference dashed line corresponds to the expected value if the contributions were uniform. **(C)** Individual scores plotted on the space defined by dimensions 1 and 2. Quantitative factors are plotted in grey arrows (here, the arrows were scaled by a factor 4 to facilitate visualization; the correlation circle for the quantitative variables and the first two dimensions is shown in supplemental material, Fig. 2). Dis, disabled patients; Int, intermediate patients; Rec, patients with good recovery.

The correlation between quantitative variables in the plane defined by the first two dimensions is illustrated by the arrows in Fig. 2C. Arrows that are close to one another indicate that the associated variables are highly positively correlated, which is consistent with results in Table 3 (e.g., MD and RD, left and right intrahemispheric functional connectivity). Arrows nearly opposed to one another indicate very high anti-correlation between variables (such as FA and RD). The longer the arrows, the more reliable their interpretation (see also the correlation circle provided in supplemental material, Fig. 2): AD and interhemispheric functional connectivity were the least reliable measures. Interestingly, the arrows for the functional connectivity metrics were nearly orthogonal to those for the structural and diffusion measures. This suggests that functional connectivity did not explain the same variability in the data, or did not convey the same kind of information, as structural and diffusion MRI.

Individual patients can also be plotted in this two-dimensional space, and this shows clear dissociation between the three groups of patients. First, let us focus on the repartition of the patients along the first dimension Dim1, which splits the space into left and right halves. All disabled patients were grouped in the left half part of the graph; they were characterized by the lowest mean splenium volumes (Fig. 1E) and the lowest mean FA measures (Fig. 1A). Most (86.4%) of the patients with good recovery were in the right half part; they had the highest mean splenium volumes and the highest mean FA measures. This shows that “Dis” and “Rec” patients essentially differed in terms of structural and diffusion metrics.

Intermediate “Int” patients had no such clear distinction in that aspect: they were mostly grouped close to the origin of the vertical axis, meaning that neither volume nor diffusion metrics reliably explained their outcome, and 61.9% of them only were on the right half side. Then, let us examine the repartition of the patients along the second dimension Dim2, which splits the space into top and bottom halves and is mainly characterized by functional connectivity measures. The majority of the intermediate “Int” patients (69.0%) were on the opposite side (here, in the top half of the graph) compared with all but one “Rec” patients, who were in the bottom half of the graph. This means that while “Int” patients could not be distinguished from other patients in terms of structural or diffusion MRI measures, they essentially differed from “Rec” patients in terms of functional connectivity. The repartition of the “Dis” patients was less clear: 50% of these patients were plotted very close to the horizontal axis, meaning that functional connectivity was not relevant to describe them.

## 4. Discussion

To our knowledge, this study is the first integrated description of chronic moderate to severe TBI patients in terms of relationship between clinical outcome and imaging metrics, namely structural changes in the corpus callosum as measured by volumetric and diffusion MRI, and functional connectivity as measured by resting-state functional MRI. A well-defined large sample (*N*=74) of patients with long-term follow-up and outcome stratification was included in this retrospective study. Imaging data were acquired on the same MRI device with the same protocol, thus avoiding biases such as small samples without differentiation in terms of severity, or acquisitions on machines from different manufacturers [31].

We resorted to multiple factorial analysis, an exploratory multivariate statistical method to summarize the data and extract relevant characteristics. Clinical outcome (GOSE score) proved to be a structuring variable that allowed us to describe the patients depending on their severity, in terms of structural and functional features.

First, multiple factorial analysis showed that structural metrics conveyed complementary information: volume loss in the corpus callosum was positively correlated with FA decrease. This enabled to distinguish between patients with good recovery (“Rec” patients), mostly clustered in the opposite part of the individual plot (Fig. 2C) compared with disabled (“Dis”) patients. This suggests that diffusion metrics and callosal volume were more altered in “Dis” than in “Rec” patients, reflecting both axonal and myelin degeneration and widespread diffuse axonal injury. Decreased FA in the splenium, as well as increased MD and AD, suggested that due to diffuse axonal injury, water diffusion was no longer restricted along the axons. This result is consistent with literature about TBI. For instance, post-acute/chronic patients with mild traumatic brain injury showed higher AD value than moderate-to-severe patients, and all patients had lower FA and higher MD values than controls, particularly in the corpus callosum [32]. The severity of diffuse axonal injury relates to outcome in patients with traumatic brain injury. For example, the extent of diffuse axonal injury in the splenium relates not only to initial injury severity, ranging from mild to severe, but also to short-term clinical outcome [33]. This may be due to the location of the splenium in the brain: The falx cerebri, the largest intracranial dura mater that separates the cerebral hemispheres, attaches to the corpus callosum at the splenium [34]. As a result, when an external force acts on the head, the splenium is limited in movement compared to the anterior portions of the corpus callosum, which makes it more susceptible to strain and injury [35]. Previous studies have also found that lesions in the genu relate to long-term disability in patients with traumatic brain injury [36].

Second, MFA showed that functional connectivity variables were orthogonal to structural variables (Fig. 2C), which means that they add complementary information to structural metrics. We indeed found that intermediate patients could not be described solely in terms of structural deficits, since they were clustered close to the origin of the first axis, suggesting that structural features were not their most relevant characteristic and that they shared structural features both with disabled patients and patients with good recovery. Interestingly, on the contrary, they were nearly all clustered in the top half of the plot of individuals, meaning that intermediate patients could be distinguished from all other patients by their functional connectivity. This is undoubtedly the most interesting finding of this study.

Here, we focused on interhemispheric and intrahemispheric connectivity between regions mediated by the corpus callosum. Median interhemispheric functional connectivity tended to be lower in intermediate patients than in patients with good recovery (Fig. 1F). As functional connectivity is thought to reflect anatomical connectivity, a more important reduction in interhemispheric functional connectivity in “Dis” patients than in “Rec” patients reasonably follows decrements in corpus callosum structural integrity. An explanation may be that sheath damage and degeneration may contribute to elevated levels of ion leakage, resulting in decreased nerve impulse propagation and diminished connectivity [37]. On the other hand, median intrahemispheric functional connectivity tended to be lower in intermediate patients than in disabled patients (Fig. 1G-H). In rhesus monkeys, sectioning of the corpus callosum led to decreased interhemispheric functional connectivity but increased intrahemispheric functional connectivity, which suggests that the loss of some structural connections may increase functional connectivity via other routes [38]. In pediatric traumatic brain injury, interhemispheric functional connectivity was reduced after corpus callosotomy, while intrahemispheric functional connections remained constant, suggesting the major role of the corpus callosum in maintaining interhemispheric coherence of spontaneous BOLD fluctuations [39]. Functional connectivity has also been recently studied in adult patients in link with cognitive performance. In a small (*N*=11) sample of patients compared with controls, a global connectivity measure (the intrinsic connectivity contrast) was significantly linked with cognitive measures such as executive function, auditory-verbal episodic memory, and cognitive reserve, but with no explicit link with the severity of the deficit after a traumatic brain injury [17]. The authors hypothesized that reduced structural connectivity and reduced brain volume on the long term following the injury resulted in reduced global functional connectivity in some brain areas but also could produce increased global functional connectivity in others as a compensation mechanism, and we provide supporting findings. To study connectivity more precisely, other studies analyzed the inter- and/or intrahemispheric functional connectivity just as we did. Interhemispheric connectivity was compared between networks in patients with TBI, showing less interhemispheric connectivity in patients than in controls especially in the executive control and fronto-parietal networks [40]. Functional connectivity between areas located in the same hemisphere was higher than between homologous regions belonging to the same resting-state network (thus cross-hemispheric), which is consistent with our findings. All resting-state networks in traumatic brain injury have also been largely studied with graph theory [41]. The connectivity between networks (default mode, dorsal attentional, fronto-parietal control networks) has been shown to decrease more than the connectivity within networks in patients compared with controls. Patterns of disruptions occurred predominantly in long-range and interhemispheric connections, which the authors attribute to damage in the corpus callosum.

None of these studies, however, attempted to distinguish between patients in terms of severity, presumably due to the small samples available. The large sample of the present study allows us to show that the interhemispheric functional connectivity in intermediate patients differed from that in recovered patients. In that respect, our results are in line with those of a recent review paper, which shows that damage to neural systems paradoxically results in enhanced functional connectivity between network regions, a phenomenon known as “hyperconnectivity” [42]. Adaptive hyperconnectivity is thought to operate as a compensatory mechanism to retain a network topology that maintains communication while minimizing the costs of network connection and maximizing network efficiency, through the recruitment of available detour paths. The observed tendency towards a higher interhemispheric connectivity in patients with a good recovery than in intermediate patients may reflect this phenomenon.

The present study is mainly limited by the fact that it is purely descriptive, showing only correlations between metrics. It would therefore be presumptuous to conclude concerning a cause-and-effect relationship between functional connectivity modifications and structural lesions. Also, no classification of the patients or prediction model could be derived from this study given the small number of disabled patients (*N*=10). We did not include neuropsychological scores and only focused on imaging metrics given the heterogeneity of the patients in terms of severity and care. This would have implied accounting for the possible confounding effect of rehabilitation and perhaps limiting the analysis to specific cognitive deficits. There are several other avenues for future exploration. This study focused on patients several years post injury. As research has indicated that neurodegeneration after a traumatic brain injury is progressive, a longitudinal study should be conducted to elucidate the changes due to diffuse axonal injuries in the corpus callosum and their relationship to functional connectivity [43]. Fiber tractography may also aid in determining the relationship between anatomical and functional connectivity. Finally, further investigations should be conducted to address whether intrahemispheric functional connectivity increase in patients with severe disability is a compensatory mechanism or a deleterious effect.

In conclusion, we showed that disabled patients had more widespread structural alterations in the corpus callosum and a lower interhemispheric connectivity than patients who made a good recovery. Most importantly, intermediate patients could be distinguished from disabled patients by a lower intrahemispheric connectivity, revealing complex long-term functional impact of moderate-to-severe traumatic brain injury.

## 5. Data availability

The data that support the findings of this study are available in Zenodo research data repository at https://doi.org/10.5281/zenodo.6641946.

## Appendix 1

Separate analyses were conducted for the genu, the body, and the splenium of the corpus callosum. Interhemispheric and intrahemispheric functional connectivities were computed as follows (see an illustration for the splenium in supplemental material, Fig. 1).

First, absolute values of correlation coefficients were converted to *Z* scores using Fisher’s *Z*-transformation [44]:

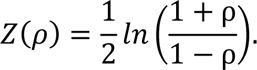

### Intrahemispheric functional connectivity

The functional connectivity FC_*w*_ between a given region *R*_*i*_ and all other regions *R*_*j*,*j*≠*i*_ in the same hemisphere, connected by the same subdivision (genu, body or splenium) of the corpus callosum, was computed as follows:

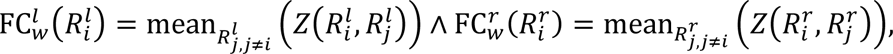

where *l* denotes the left hemisphere and *r* denotes the right hemisphere. Finally, the left intrahemispheric connectivity 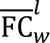 and the right intrahemispheric connectivity 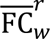, for regions connected by the same subdivision of the corpus callosum, were computed as follows:

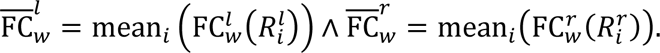

### Interhemispheric functional connectivity

The functional connectivity 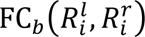 between two homologous regions 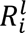 in the left hemisphere and 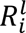 in the right hemisphere was computed as follows:

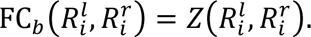

The interhemispheric connectivity 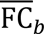 for all regions connected by a given subdivision of the corpus callosum (genu, body or splenium) was then defined as the average of all FC_b_ between homologous regions connected by the same subdivision:

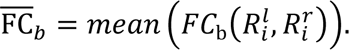

## Appendix 2

Multiple factorial analysis (MFA) is a multivariate data analysis method that summarizes a complex data table in which individuals (here, the patients) are described by several sets of variables, which can be quantitative (here, imaging variables, which are scaled to unit variance) and/or categorical (here, GOSE category: disabled patients, intermediate patients, or patients who have made a good recovery) structured into groups. To balance the influence of each set of variables, variables in the same group are normalized using the same weighting value, which can vary from one group to another. MFA then takes the contribution of all active groups of variables into account to define the distance between individuals. Roughly, this method relies on principal component analysis when for quantitative variables, and multiple correspondence analysis when variables are qualitative.

## Abbreviations

AAL: Automated Anatomical Labeling
AD: axial diffusivity
ANOVA: analysis of variance
BOLD: blood-oxygen-level dependent
DTI: diffusion tensor imaging
FA: fractional anisotropy
GCS: Glasgow Coma Scale
GOSE: Glasgow Outcome Scale extended
MANOVA: multivariate ANOVA
MD: mean diffusivity
MFA: multiple factorial analysis
MRI: magnetic resonance imaging
RD: radial diffusivity
rs-fMRI: resting-state functional
MRI TBI: traumatic brain injury

