## supplemental material for "Multimodal magnetic resonance imaging characterizes clinical outcome in chronic traumatic brain injury"

### MRI data acquisition

Whole-brain MRI acquisitions were performed on a 3.0 Tesla GE Medical Systems (Milwaukee, WI) SIGNA scanner in the Neuroradiology Department of the Pitié-Salpêtrière Hospital (Paris, France), using a standard 8-channel head coil (8HRBRAIN), in the following order.

Diffusion-weighted images were acquired using a single-shell echo-planar imaging (EPI) sequence (50 gradient-encoded directions, b-value=1000 s/mm², 50 contiguous axial slices per volume, 128x128 matrix, voxel size 2x2x2.5 mm, TR/TE: 14000/85 ms, flip angle 90°, A/P phase encoding). One volume ($b_0$) was acquired without use of a diffusion gradient (b = 0 s/mm$^2$). No cardiac gating was used.

High-resolution 3D $T_1$-weighted acquisition consisted of an inversion-recovery MPRAGE pulse sequence (192 sagittal slices, 256x256 matrix, 1 mm isotropic voxels, TR/TE/TI: 7.192/3.104/380 ms, flip angle 15°).

Resting-state fMRI (rs-fMRI) acquisition consisted of a gradient-echo EPI sequence (200 repetitions, 50 contiguous axial slices per volume, 64x64 matrix, voxel size 3.5x3.5x3 mm, TR/TE: 2400/30 ms, flip angle 90°, interleaved acquisition) during which participants were instructed to remain still, stay awake, keep their eyes open, and refrain from any overt activity. Run duration was 8 minutes.

### MRI data preprocessing

#### Structural data preprocessing

Individual 3D $T_1$-weighted structural images were preprocessed using the Freesurfer image analysis suite[†], which included motion and bias correction of the $T_1$-weighted image, removal of non-brain tissue, and segmentation of the subcortical white matter and deep gray matter volumetric structures [1].

#### DTI data preprocessing

DTI data was motion-corrected by linearly registering the gradient-encoded volumes to the $b_0$ volume. The diffusion tensor was estimated in each voxel using an ordinary least-squares approach from the FSL software [2][‡], leading to four parametric maps: fractional anisotropy (FA), mean diffusivity (MD), diffusion along the main axis (axial diffusivity AD), and diffusion along the transverse plane (radial diffusivity RD) [3]. The four maps were then registered to the MNI standard space using NiftyReg toolbox [4][#], by combining (1) an affine transformation from the FA map to the individual 3D $T_1$-weighted image in the subject's space and (2) an affine followed by a nonlinear transformation from the subject's $T_1$-weighted image to the MNI space.

#### Resting-state fMRI data preprocessing

Functional MRI time series were motion corrected using rigid body alignment (FSL[‡]) and registered to the MNI standard space by combining (1) an affine transformation from the mean functional volume to the individual 3D $T_1$-weighted image in the subject's space and (2) an affine followed by a nonlinear transformation from the subject's $T_1$-weighted image to the MNI space (NiftyReg[#]). Very low-frequency drifts were removed by high-pass filtering (cut-off frequency: 0.005 Hz), physiological noise correction was performed using CompCor and a band-pass filter of [0.005-1.0 Hz] was applied [5].

---

[†] surfer.nmr.mgh.harvard.edu, version number: 5.3.0

[‡] www.fmrib.ox.ac.uk/fsl, version number: 5.0.9

[#] cmictig.cs.ucl.ac.uk/wiki/index.php/NiftyReg,
Git revision number 83d8d1182ed4c227ce4764f1fdab3b1797eecd8d

#### Interhemispheric and intrahemispheric functional connectivity

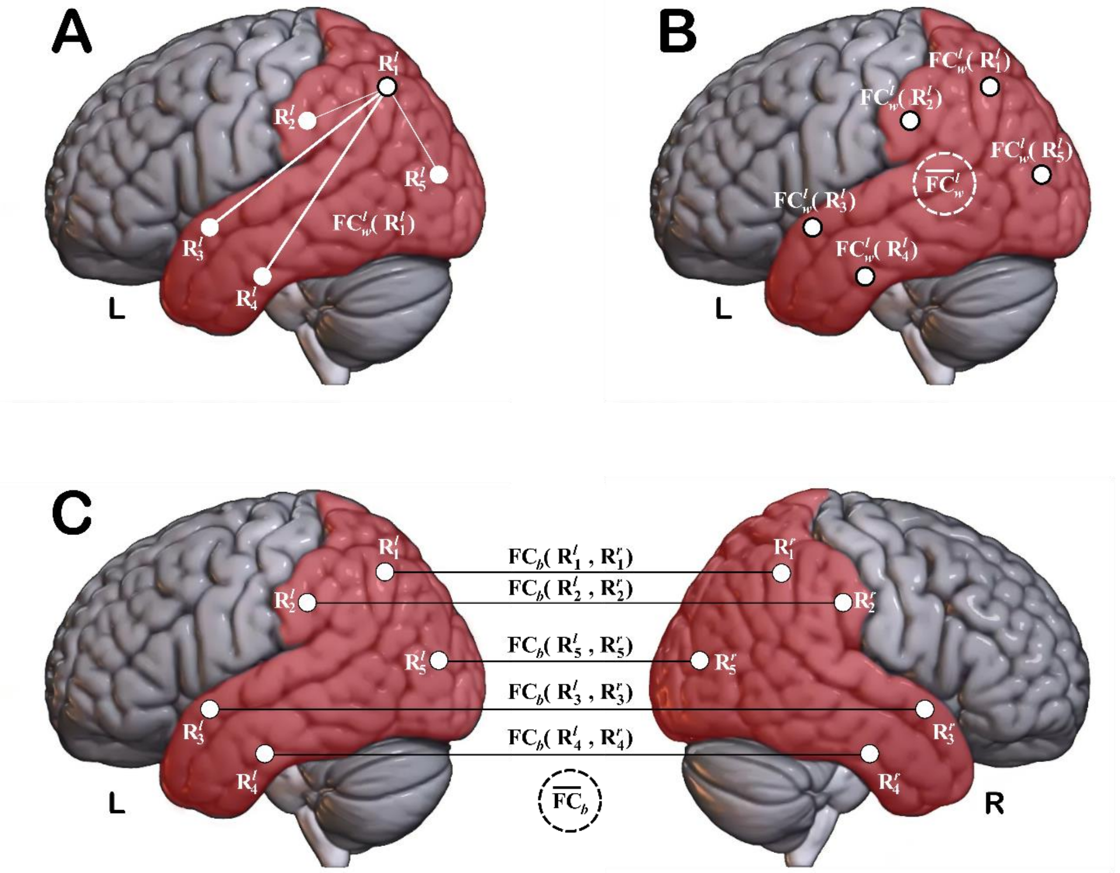

**Figure 1.** Illustration of how functional connectivity is computed for regions connected by the splenium of the corpus callosum. *Intrahemispheric connectivity*: **(A)** the functional connectivity $\text{FC}_w^l(R_1^l)$ between a given region $R_1$ and all other regions $R_2$, $R_3$, $R_4$ and $R_5$ in the left hemisphere is computed; **(B)** the final intrahemispheric connectivity $\overline{FC}_w^l$ for the left hemisphere is obtained by averaging all $\text{FC}_w^l(R_i^l)$ values. *Interhemispheric connectivity*: **(C)** all connectivity values $\text{FC}_b(R_i^l, R_i^r)$ between homologous pairs of regions ($R_i^l$ in the left hemisphere and $R_i^r$ in the right hemisphere) are computed, and averaged to yield the final interhemispheric connectivity $\overline{FC}_b$.

### Detailed patient characteristics

**Table 1.** Age range is at the time of GOSE assessment. M, male; F, female; GCS, Glasgow Coma Scale score (unavailable for three patients); GOSE, Glasgow Outcome Scale Extended score.

| Patient | Age (in years) | Gender | Mechanism of injury | Initial GCS | GOSE | Delay to GOSE assessment (in months) | Delay to MRI (in months) |
|---|---|---|---|---|---|---|---|
| P01 | 25-30 | F | Road traffic accident | 3 | 3 | 54 | 55 |
| P02 | > 50 | M | Road traffic accident | 12 | 3 | 61 | 61 |
| P03 | 25-30 | M | Road traffic accident | 5 | 3 | 83 | 89 |
| P04 | 31-35 | M | Road traffic accident | 5 | 4 | 37 | 37 |
| P05 | 25-30 | M | Road traffic accident | 6 | 4 | 41 | 41 |
| P06 | 36-40 | M | Road traffic accident | 6 | 4 | 46 | 46 |
| P07 | 25-30 | M | Road traffic accident | 6 | 4 | 55 | 55 |
| P08 | 31-35 | M | Assault | 7 | 4 | 70 | 69 |
| P09 | 41-50 | M | Assault | 6 | 4 | 84 | 84 |
| P10 | 41-50 | M | Road traffic accident | 3 | 4 | 106 | 106 |
| P11 | 25-30 | M | Other | 8 | 5 | 31 | 31 |
| P12 | 18-24 | M | Road traffic accident | 8 | 5 | 34 | 33 |
| P13 | 18-24 | F | Road traffic accident | 7 | 5 | 36 | 36 |
| P14 | > 50 | M | Road traffic accident | 9 | 5 | 39 | 39 |
| P15 | 18-24 | F | Road traffic accident | 6 | 5 | 39 | 39 |
| P16 | 31-35 | M | Road traffic accident | 3 | 5 | 41 | 41 |
| P17 | 36-40 | M | Road traffic accident | 3 | 5 | 43 | 43 |
| P18 | 36-40 | M | Assault | 8 | 5 | 45 | 45 |
| P19 | > 50 | M | Fall | 8 | 5 | 46 | 46 |
| P20 | 41-50 | M | Road traffic accident | 8 | 5 | 46 | 46 |
| P21 | 18-24 | M | Other | 6 | 5 | 47 | 47 |
| P22 | 41-50 | M | Assault | 6 | 5 | 47 | 48 |
| P23 | 25-30 | M | Assault | 3 | 5 | 57 | 57 |
| P24 | > 50 | M | Road traffic accident | 8 | 5 | 58 | 58 |
| P25 | > 50 | M | Other | 7 | 5 | 60 | 61 |
| P26 | 25-30 | M | Fall | 5 | 5 | 60 | 61 |
| P27 | 36-40 | M | Road traffic accident | 3 | 5 | 62 | 62 |
| P28 | 41-50 | M | Road traffic accident | 3 | 5 | 64 | 64 |
| P29 | 31-35 | M | Road traffic accident | 3 | 5 | 83 | 84 |
| P30 | 41-50 | M | Road traffic accident | 3 | 5 | 78 | 91 |
| P31 | > 50 | M | Fall | - | 5 | 92 | 93 |
| P32 | 41-50 | M | Road traffic accident | 6 | 5 | 119 | 117 |
| P33 | 18-24 | F | Road traffic accident | 7 | 6 | 34 | 34 |
| P34 | 18-24 | M | Road traffic accident | 4 | 6 | 35 | 38 |
| P35 | 18-24 | M | Road traffic accident | 6 | 6 | 42 | 42 |
| P36 | 25-30 | M | Fall | 12 | 6 | 45 | 45 |
| P37 | 36-40 | M | Road traffic accident | 3 | 6 | 51 | 51 |
| P38 | 18-24 | M | Road traffic accident | 6 | 6 | 48 | 51 |
| P39 | 31-35 | M | Road traffic accident | 8 | 6 | 53 | 54 |
| P40 | 25-30 | M | Other | 8 | 6 | 63 | 63 |
| P41 | > 50 | M | Road traffic accident | 8 | 6 | 63 | 63 |
| P42 | 31-35 | M | Fall | 7 | 6 | 66 | 69 |
| P43 | 25-30 | M | Assault | 3 | 6 | 74 | 74 |
| P44 | > 50 | M | Fall | 4 | 6 | 75 | 76 |
| P45 | 41-50 | M | Road traffic accident | 8 | 6 | 78 | 78 |
| P46 | 41-50 | M | Fall | 7 | 6 | 82 | 82 |
| P47 | 25-30 | M | Road traffic accident | 11 | 6 | 83 | 82 |
| P48 | 25-30 | M | Road traffic accident | 8 | 6 | 84 | 84 |
| P49 | 36-40 | M | Road traffic accident | 8 | 6 | 86 | 86 |
| P50 | > 50 | M | Fall | 4 | 6 | 95 | 94 |
| P51 | 25-30 | M | Fall | - | 6 | 94 | 94 |
| P52 | > 50 | M | Road traffic accident | 8 | 6 | 98 | 98 |
| P53 | 18-24 | M | Road traffic accident | 3 | 7 | 32 | 33 |

| | | | | | | | |
|---|---|---|---|---|---|---|---|
| P54 | 31-35 | F | Road traffic accident | 8 | 7 | 34 | 34 |
| P55 | 18-24 | M | Road traffic accident | 6 | 7 | 42 | 42 |
| P56 | 31-35 | M | Assault | 8 | 7 | 43 | 43 |
| P57 | 25-30 | F | Road traffic accident | 7 | 7 | 43 | 44 |
| P58 | > 50 | M | Road traffic accident | 3 | 7 | 47 | 47 |
| P59 | 18-24 | M | Fall | 9 | 7 | 44 | 50 |
| P60 | > 50 | M | Fall | 12 | 7 | 54 | 54 |
| P61 | 31-35 | M | Other | 12 | 7 | 62 | 63 |
| P62 | 36-40 | M | Road traffic accident | 10 | 7 | 76 | 76 |
| P63 | 18-24 | M | Assault | - | 7 | 76 | 76 |
| P64 | 31-35 | M | Road traffic accident | 9 | 7 | 77 | 77 |
| P65 | > 50 | F | Fall | 13 | 7 | 84 | 83 |
| P66 | > 50 | M | Assault | 9 | 7 | 93 | 93 |
| P67 | 25-30 | M | Road traffic accident | 6 | 7 | 95 | 95 |
| P68 | 36-40 | M | Road traffic accident | 11 | 7 | 113 | 113 |
| P69 | 25-30 | M | Road traffic accident | 5 | 8 | 39 | 38 |
| P70 | 25-30 | M | Assault | 3 | 8 | 45 | 45 |
| P71 | 18-24 | M | Other | 7 | 8 | 54 | 53 |
| P72 | 41-50 | F | Road traffic accident | 11 | 8 | 70 | 70 |
| P73 | > 50 | M | Road traffic accident | 3 | 8 | 76 | 80 |
| P74 | > 50 | M | Road traffic accident | 10 | 8 | 83 | 83 |

### Results for the splenium of the corpus callosum

**Figure 2.** Results of the multivariate factorial analysis for the splenium of the corpus callosum. Correlation between the quantitative variables and the first two dimensions Dim1 (explaining 30.8% of the total inertia) and Dim2 (explaining 17.8% of the total inertia).

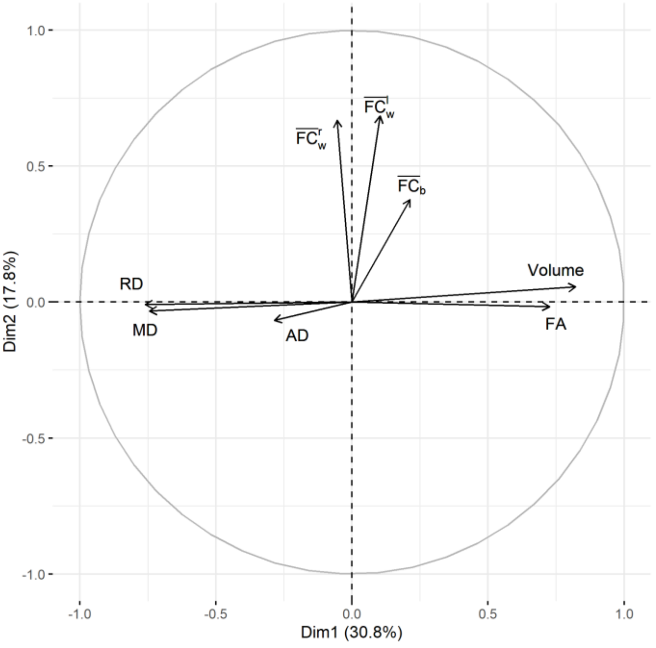

### Results for the body of the corpus callosum

**Figure 3.** Distribution of imaging metrics for the body of the corpus callosum. Dis, disabled patients; Int, intermediate patients; Rec, patients with good recovery. Black dots: mean values, vertical black lines: standard deviation, grey dots: individual values. Panels **A**: fractional anisotropy (FA), **B**: mean diffusivity (MD), **C**: axial diffusivity (AD), **D**: radial diffusivity (RD), **E**: volume, **F**: interhemispheric functional connectivity ($\overline{\mathrm{FC}}_b$), **G-H**: left ($\overline{\mathrm{FC}}_w^l$) and right ($\overline{\mathrm{FC}}_w^r$) intrahemispheric functional connectivity.

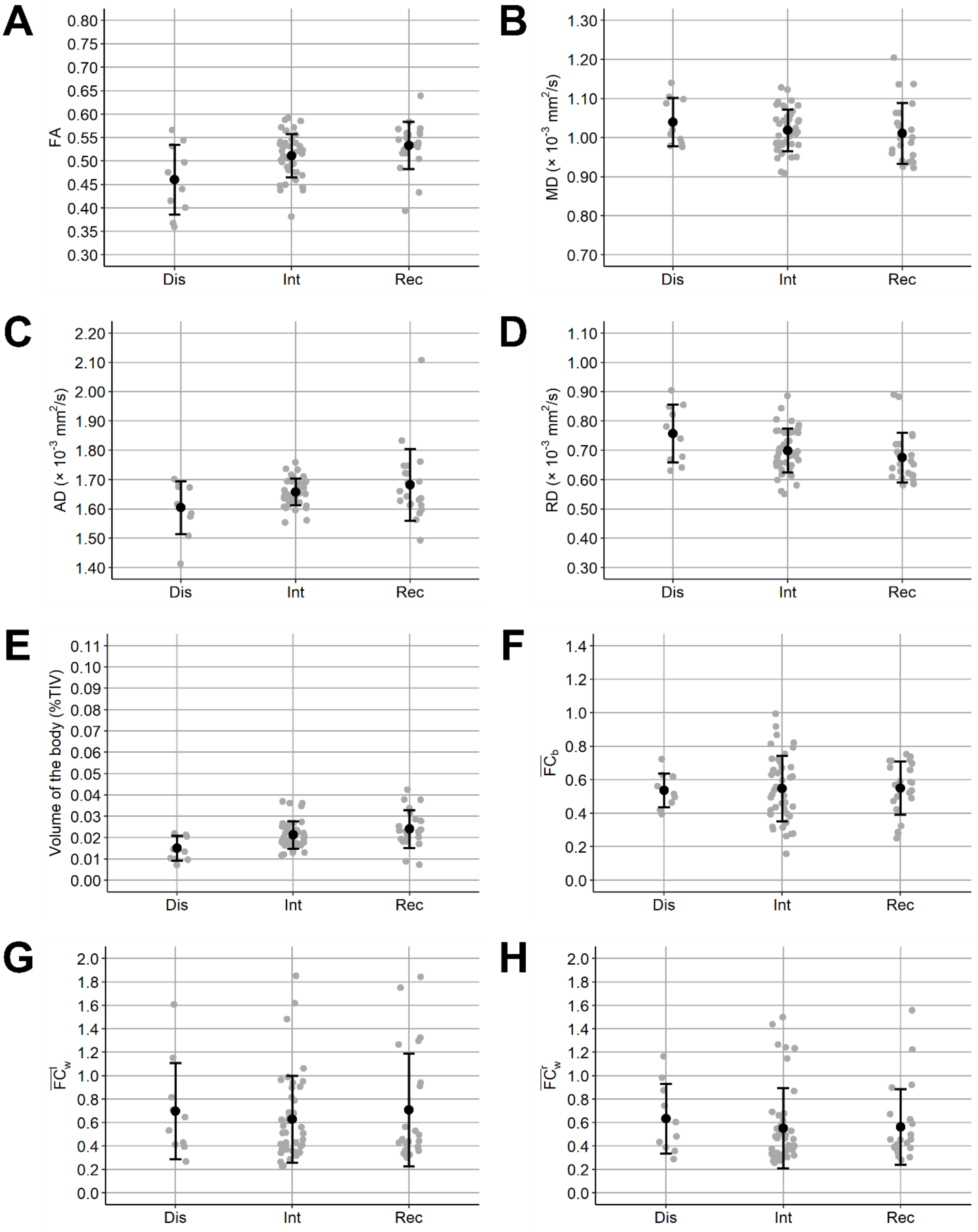

**Table 2.** Mean [standard deviation] of the imaging metrics, for the body of the corpus callosum, in each group of patients. Dis, disabled patients; Int, intermediate patients; Rec, patients with good recovery. TIV: total intracranial volume, FA: fractional anisotropy, MD: mean diffusivity, AD: axial diffusivity, RD: radial diffusivity, $\overline{\mathrm{FC}}_b$: interhemispheric functional connectivity, $\overline{\mathrm{FC}}_w^l$: left and $\overline{\mathrm{FC}}_w^r$: right intrahemispheric functional connectivity.

| | Volume (%TIV) | FA | MD ($x10^{-3}$ $mm^2/s$) | AD ($x10^{-3}$ $mm^2/s$) | RD ($x10^{-3}$ $mm^2/s$) |
|---|---|---|---|---|---|
| Dis | 0.015 [0.006] | 0.460 [0.074] | 1.04 [0.061] | 1.60 [0.091] | 0.757 [0.099] |
| Int | 0.021 [0.006] | 0.511 [0.047] | 1.02 [0.053] | 1.66 [0.046] | 0.699 [0.075] |
| Rec | 0.024 [0.009] | 0.533 [0.050] | 1.01 [0.078] | 1.68 [0.122] | 0.675 [0.085] |
| | $\overline{\mathrm{FC}}_b$ | $\overline{\mathrm{FC}}_w^l$ | $\overline{\mathrm{FC}}_w^r$ | | |
| Dis | 0.536 [0.101] | 0.697 [0.410] | 0.633 [0.297] | | |
| Int | 0.548 [0.196] | 0.628 [0.371] | 0.551 [0.342] | | |
| Rec | 0.158 [0.550] | 0.708 [0.482] | 0.563 [0.322] | | |

**Table 3.** Spearman's rank correlation matrix between MRI measures. Correlations indicated in bold are significantly different from 0 (*P*<0.05 Bonferroni-corrected for 28 different pairwise tests, i.e. *P*<0.0018).

| | Volume | FA | MD | AD | RD | $\overline{\mathrm{FC}}_b$ | $\overline{\mathrm{FC}}_w^l$ |
|---|---|---|---|---|---|---|---|
| FA | **0.63** | - | - | - | - | - | - |
| MD | **-0.58** | **-0.73** | - | - | - | - | - |
| AD | 0.02 | 0.15 | **0.45** | - | - | - | - |
| RD | **-0.68** | **-0.94** | **0.91** | 0.13 | - | - | - |
| $\overline{\mathrm{FC}}_b$ | 0.04 | 0.01 | -0.04 | -0.04 | -0.02 | - | - |
| $\overline{\mathrm{FC}}_w^l$ | 0.12 | 0.02 | 0.07 | 0.08 | 0.01 | 0.29 | - |
| $\overline{\mathrm{FC}}_w^r$ | 0.02 | -0.04 | 0.12 | 0.02 | 0.06 | **0.36** | **0.75** |

Permutational MANOVA was conducted with body volume, FA and AD as independent variables, showing a significant group effect (*F*=7.20, *P*=0.002). Body volume was found to differ significantly (Kruskal-Wallis $\chi^2$=9.70, *P*=0.008) between "Dis" and "Rec" patients (Dunn's test *Z*=3.11, *P*=0.01) and between "Dis" and "Int" patients (Dunn's test *Z*=2.28, *P*=0.04). FA was found to differ significantly (Kruskal-Wallis $\chi^2$=8.74, *P*=0.01) between "Dis" and "Rec" patients (Dunn's test *Z*=2.89, *P*=0.01).

A separate permutational MANOVA was conducted with connectivity measures as independent variables, showing no significant difference between patient groups (*F*=0.25, *P*=0.90).

**Figure 4.** Results of the multivariate factorial analysis for the body of the corpus callosum. Correlation between the quantitative variables and the first two dimensions Dim1 (explaining 30.8% of the total inertia) and Dim2 (explaining 17.4% of the total inertia).

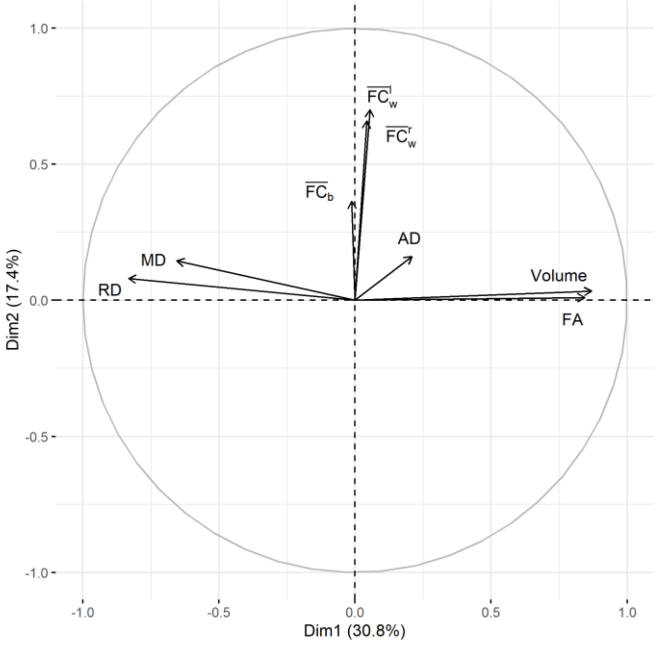

**Figure 5.** Results of the multivariate factorial analysis for the body of the corpus callosum. **(A-B)**: contribution (in %) of each group of variables to the first two dimensions. The reference dashed line corresponds to the expected value if the contributions were uniform. **(C)**: Individual scores plotted on the space defined by dimensions 1 and 2. Quantitative factors are plotted in grey arrows (here, the arrows were scaled by a factor 4 to facilitate visualization). Dis, disabled patients; Int, intermediate patients; Rec, patients with good recovery.

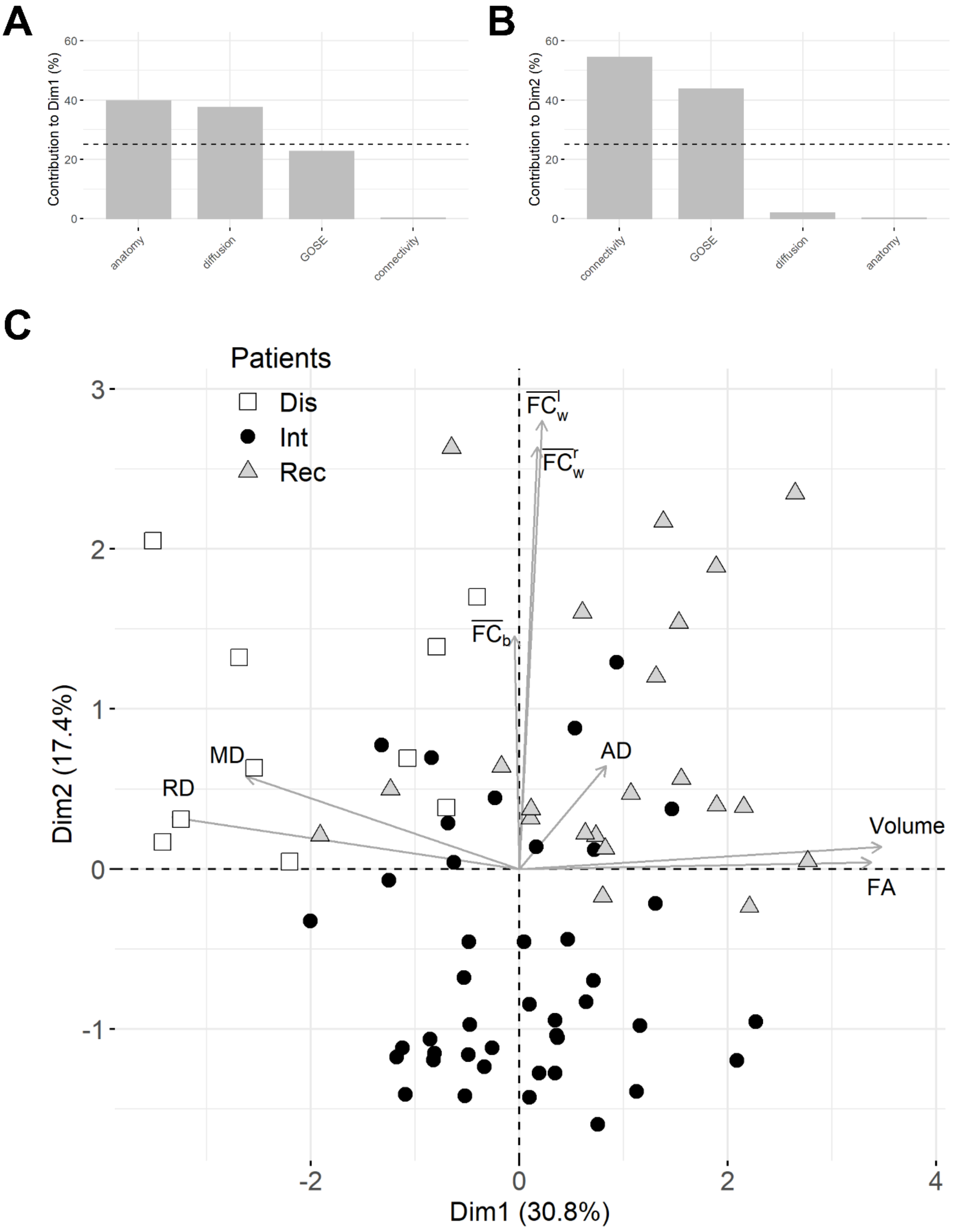

### Results for the genu of the corpus callosum

**Figure 6.** Distribution of imaging metrics for the genu of the corpus callosum. Dis, disabled patients; Int, intermediate patients; Rec, patients with good recovery. Black dots: mean values, vertical black lines: standard deviation, grey dots: individual values. Panels **A**: fractional anisotropy (FA), **B**: mean diffusivity (MD), **C**: axial diffusivity (AD), **D**: radial diffusivity (RD), **E**: volume, **F**: interhemispheric functional connectivity ($\overline{\mathrm{FC}}_b$), **G-H**: left ($\overline{\mathrm{FC}}_w^l$) and right ($\overline{\mathrm{FC}}_w^r$) intrahemispheric functional connectivity.

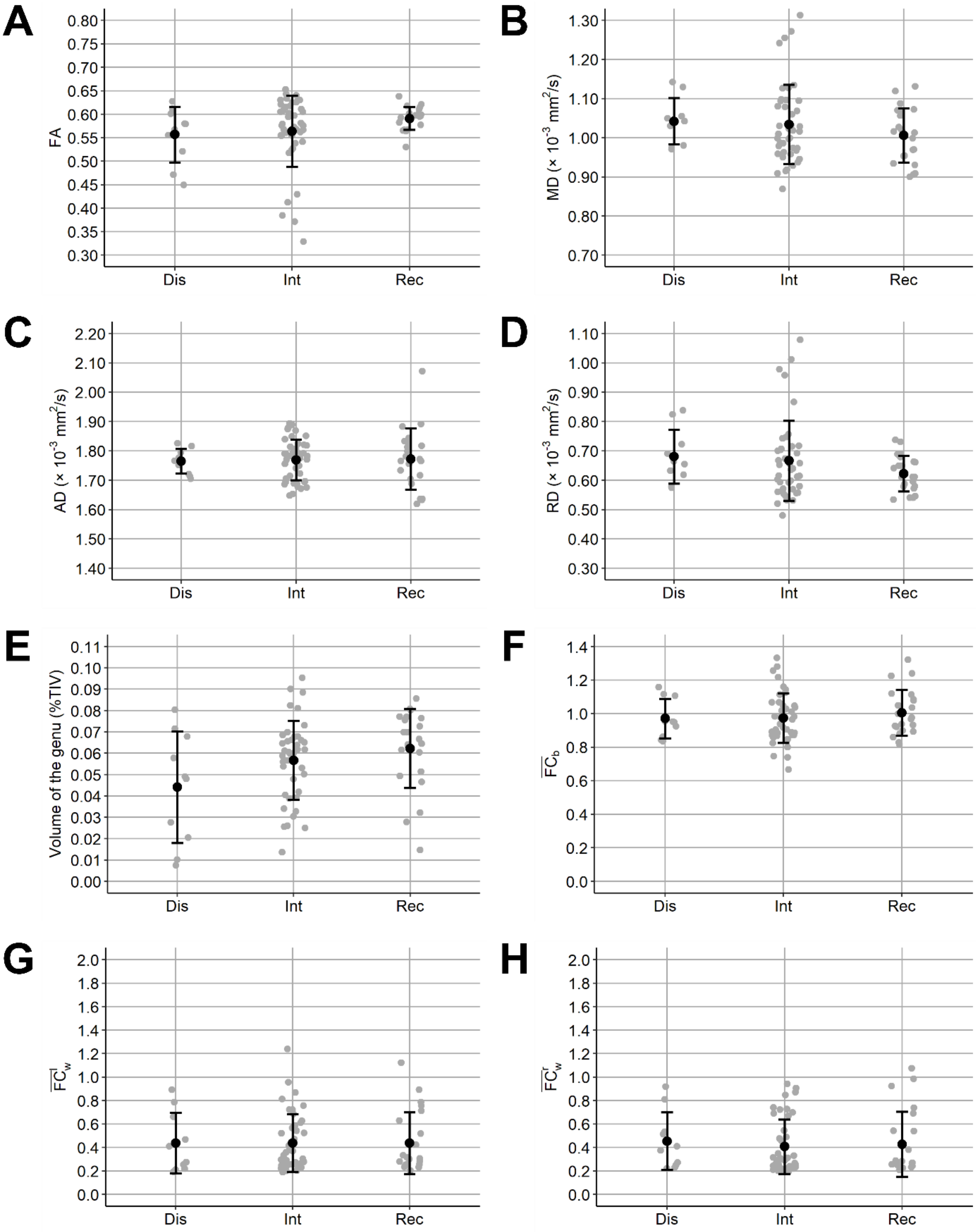

**Table 4.** Mean and standard deviation (SD) of the imaging metrics, for the genu of the corpus callosum, in each group of patients. Dis, disabled patients; Int, intermediate patients; Rec, patients with good recovery. TIV: total intracranial volume, FA: fractional anisotropy, MD: mean diffusivity, AD: axial diffusivity, RD: radial diffusivity, $\overline{\mathrm{FC}}_b$: interhemispheric functional connectivity, $\overline{\mathrm{FC}}_w^l$: left and $\overline{\mathrm{FC}}_w^r$: right intrahemispheric functional connectivity.

| | Volume (%TIV) | FA | MD ($x10^{-3}$ $mm^2/s$) | AD ($x10^{-3}$ $mm^2/s$) | RD ($x10^{-3}$ $mm^2/s$) |
|---|---|---|---|---|---|
| Dis | 0.044 [0.026] | 0.557 [0.059] | 1.04 [0.059] | 1.77 [0.042] | 0.681 [0.092] |
| Int | 0.057 [0.019] | 0.564 [0.076] | 1.03 [0.101] | 1.77 [0.070] | 0.666 [0.136] |
| Rec | 0.062 [0.019] | 0.591 [0.024] | 1.01 [0.069] | 1.77 [0.105] | 0.623 [0.061] |

| | $\overline{\mathrm{FC}}_b$ | $\overline{\mathrm{FC}}_w^l$ | $\overline{\mathrm{FC}}_w^r$ |
|---|---|---|---|
| Dis | 0.971 [0.117] | 0.439 [0.258] | 0.455 [0.245] |
| Int | 0.973 [0.148] | 0.438 [0.245] | 0.408 [0.231] |
| Rec | 1.01 [0.137] | 0.438 [0.261] | 0.428 [0.277] |

**Table 5.** Spearman's rank correlation matrix between MRI measures. Correlations indicated in bold are significantly different from 0 (*P*<0.05 Bonferroni-corrected for 28 different pairwise tests, i.e. *P*<0.0018).

| | Volume | FA | MD | AD | RD | $\overline{\mathrm{FC}}_b$ | $\overline{\mathrm{FC}}_w^l$ |
|---|---|---|---|---|---|---|---|
| FA | **0.36** | - | - | - | - | - | - |
| MD | **-0.54** | **-0.74** | - | - | - | - | - |
| AD | -0.34 | -0.15 | **0.69** | - | - | - | - |
| RD | -**0.50** | **-0.89** | **0.96** | **0.51** | - | - | - |
| $\overline{\mathrm{FC}}_b$ | -0.23 | -0.03 | -0.01 | 0.02 | 0.01 | - | - |
| $\overline{\mathrm{FC}}_w^l$ | -0.03 | -0.07 | 0.03 | 0.04 | 0.05 | 0.30 | - |
| $\overline{\mathrm{FC}}_w^r$ | -0.06 | -0.10 | 0.13 | 0.11 | 0.13 | 0.32 | **0.77** |

Permutational MANOVA was conducted with genu volume, FA and AD as independent variables, showing a significant group effect (*F*=3.01, *P*=0.03). Volume only tended to differ (Kruskal-Wallis $\chi^2$=4.94, *P*=0.08) between "Dis" and "Rec" patients (Dunn's test *Z*=-2.13, *P*=0.10).

A separate permutational MANOVA was conducted with connectivity measures as independent variables, showing no significant difference between patient groups (*F*=0.03, *P*=0.98).

**Figure 7.** Results of the multivariate factorial analysis for the genu of the corpus callosum. Correlation between the quantitative variables and the first two dimensions Dim1 (explaining 27.5% of the total inertia) and Dim2 (explaining 17.6% of the total inertia).

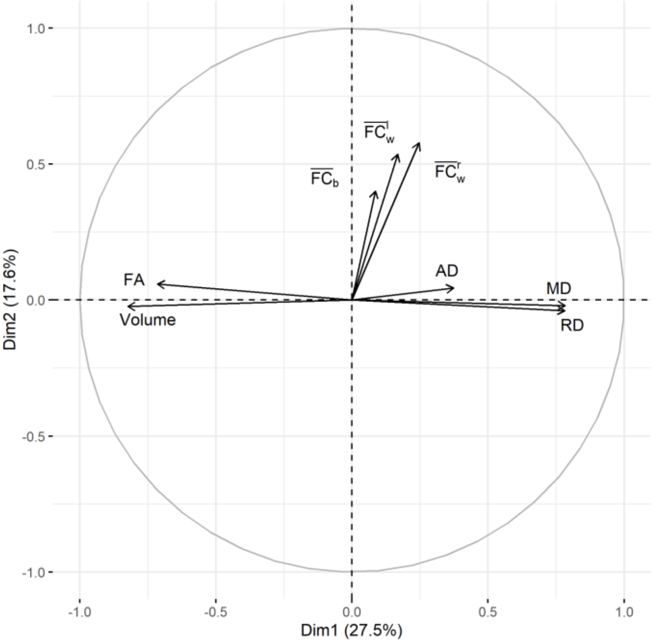

**Figure 8.** Results of the multivariate factorial analysis for the genu of the corpus callosum. **(A-B)**: contribution (in %) of each group of variables to the first two dimensions. The reference dashed line corresponds to the expected value if the contributions were uniform. **(C)**: Individual scores plotted on the space defined by dimensions 1 and 2. Quantitative factors are plotted in grey arrows (here, the arrows were scaled by a factor 4 to facilitate visualization). Dis, disabled patients; Int, intermediate patients; Rec, patients with good recovery.

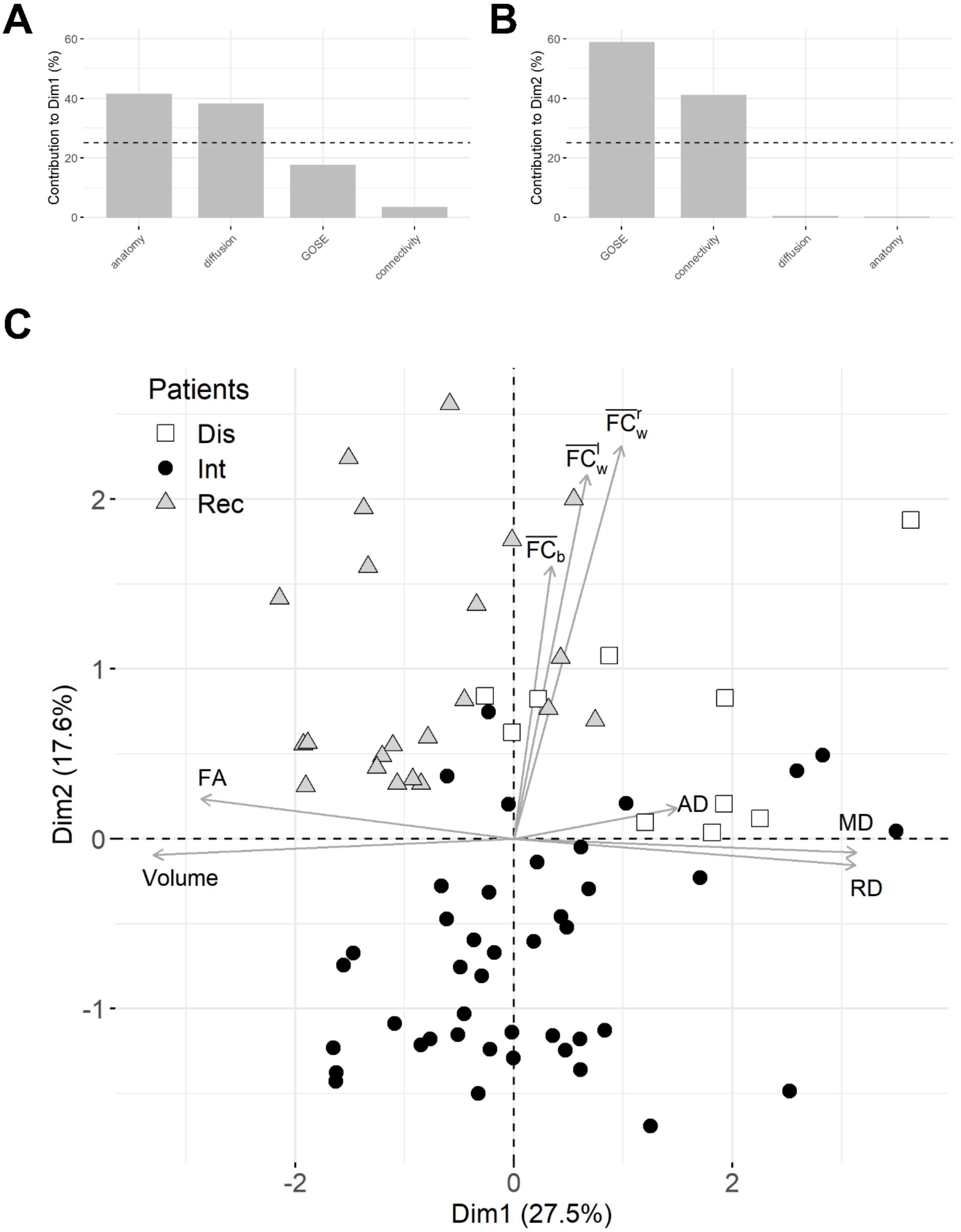
